# A simple SEIR-V model to estimate COVID-19 prevalence and predict SARS-CoV-2 transmission using wastewater-based surveillance data

**DOI:** 10.1101/2022.07.17.22277721

**Authors:** Tin Phan, Samantha Brozak, Bruce Pell, Anna Gitter, Kristina D. Mena, Yang Kuang, Fuqing Wu

**Affiliations:** Theoretical Biology and Biophysics Group, Los Alamos National Laboratory, New Mexico, USA; School of Mathematical and Statistical Sciences, Arizona State University, Arizona, USA; Department of Mathematics and Computer Science, Lawrence Technological University, MI USA; The University of Texas Health Science Center at Houston, School of Public Health Houston, Texas, USA 77030

## Abstract

Wastewater-based surveillance (WBS) has been widely used as a public health tool to monitor SARS-CoV-2 transmission. However, epidemiological inference from WBS data remains understudied and limits its application. In this study, we have established a quantitative framework to estimate COVID-19 prevalence and predict SARS-CoV-2 transmission through integrating WBS data into an SEIR-V model. We conceptually divide the individual-level viral shedding course into exposed, infectious, and recovery phases as an analogy to the compartments in population-level SEIR model. We demonstrated that the temperature effect on viral losses in the sewer can be straightforwardly incorporated in our framework. Using WBS data from the second wave of the pandemic (Oct 02, 2020 – Jan 25, 2021) in the Great Boston area, we showed that the SEIR-V model successfully recapitulates the temporal dynamics of viral load in wastewater and predicts the true number of cases peaked earlier and higher than the number of reported cases by 16 days and 8.6 folds (*R* = 0.93), respectively. This work showcases a simple, yet effective method to bridge WBS and quantitative epidemiological modeling to estimate the prevalence and transmission of SARS-CoV-2 in the sewershed, which could facilitate the application of wastewater surveillance of infectious diseases for epidemiological inference and inform public health actions.

## 1. Introduction

Wastewater-based surveillance (WBS) has been used as a public health tool to monitor SARS-CoV-2 infection in the population since the beginning of the COVID-19 pandemic. So far, WBS has been widely implemented in over 67 countries (Naughton et al., 2021). The Centers for Disease Control and Prevention (CDC) also launched the National Wastewater Surveillance System in late 2020 to monitor the spread of COVID-19 in the United States (CDC, 2020). Wastewater collates SARS-CoV-2 particles excreted by infected individuals irrespective of clinical symptoms or presentation, which provides an opportunity to capture the viral shedding prior to symptoms and estimate the true magnitude of viral infections in communities (Bivins et al., 2020b; Hart and Halden, 2020; Peccia et al., 2020; Randazzo et al., 2020; Saguti et al., 2021; Wu et al., 2022b). Previous work has shown that SARS-CoV-2 concentrations in wastewater were much higher than expected from clinically reported cases and predicted clinical reported data for 4-10 days (Wu et al., 2020, 2022b, Peccia et al., 2020), and up to 14 days (Krivoňáková et al., 2021; Karthikeyan et al. 2020). Furthermore, the fast turnaround time of wastewater and flexible sampling strategy enable WBS to provide a near real-time monitoring of the viral transmission in the sewershed. Finally, WBS is less resource intensive than the large-scale, individual-based clinical testing and thus can be used as a cost-efficient tool for monitor the trend of viral infection in the population and new variants when combined with next-generation sequencing (Bivins et al., 2020b; Safford et al., 2022; Wu et al., 2022a). These properties make WBS a feasible public health tool to monitor SARS-CoV-2 in an endemic, which can also be customized for future pandemics.

WBS has enabled researchers to estimate the total viral load in a sewershed; however, there are still limitations regarding quantifying and predicting viral transmission in a community. Few recent studies have tried to build classical susceptible-infected-removed (SIR)-type models to bridge the measured viral concentration and reported case number. For example, Proverbio et al. (2022) added a variable that keeps track of actively shedding individuals in a stochastic susceptible-exposed-infectious-recovered (SEIR) model and used a constant viral shedding rate to connect the number of infected cases to viral concentration in wastewater (Proverbio et al., 2022). Conversely, Brouwer et al. (2022) accounted for time dependent viral shedding rates by incorporating multiple subclasses with different shedding rates within each infected stage of the model to better predict viral concentrations and reported cases (Brouwer et al., 2022). A similar approach is conducted by Nourbakhsh et al. (2022), but with more sub-classification of the infected class (Nourbakhsh et al., 2022). These modeling approaches allow the modelers to connect viral concentrations in wastewater with the reported cases and predict the course of the pandemic.

Dynamical models in epidemiology thus far often overlook the opportunity to utilize biologically interpretable and experimentally measurable parameters in the link between infected people and the shed viral RNA in wastewater. The model structure is usually complicated with many parameters, so it is difficult to fully parametrize the models without running into issues such as model identifiability. Hence, our primary objective in this work is to leverage our understanding of the biology of SARS-CoV-2 shedding to construct a simple, mechanistic, dynamic model that connects viral load in wastewater with the total number of infected cases in the sewershed. Our secondary objective is to introduce the effect of wastewater temperature into the modeling framework due to its significant impact on the viral loss (or decay) rate in the sewer (Hart and Halden, 2020).

## 2. Materials and methods

### 2.1. Samples and wastewater data

Raw, 24-hour composite wastewater samples were collected from the Deer Island wastewater treatment plant in Massachusetts from October 02, 2020 to January 25, 2021. The Massachusetts wastewater treatment plant where we obtained samples has two major influent streams, which are referred to as the “northern” and “southern” influents. The daily flow rates during the sampling period for the northern and southern influents are 4.54e5 – 2.3e6 *m*^3^/*day*, and 2.16e5 – 1.19e6*m*^3^/*day*, respectively. Together the two catchments represent approximately 2.3 million wastewater customers in Middlesex, Norfolk, and Suffolk counties, primarily in urban and suburban neighborhoods. There are 5,100 miles of local sewers transporting wastewater into 227 miles of interceptor pipes to the wastewater treatment plant (www.mwra.com), and the typical turnaround time for the plant to treat wastewater is 24 hours. Samples were processed as they were received. Experimental method and data were reported in our previous work (Wu et al., 2022b; Xiao et al., 2022). Briefly, the samples were pasteurized at 60°C for 1 hour for disinfection, and then filtered with 0.2 µm hydrophilic polyethersulfone membrane (Millipore Sigma) to remove bacterial cells and debris. Then, 15-ml filtrate was concentrated to ∼200 ul with Amicon Ultra Centrifugal Filter (30-kDa cutoff, Millipore Sigma), and lysed with Qiagen AVL buffer followed by RNA extraction with Qiagen RNeasy kit. SARS-CoV-2 concentrations were quantified by one-step reverse transcription-polymerase chain reaction (RT-PCR) with the Taqman Fast Virus 1-Step Master Mix (Thermofisher) and CDC N1 and N2 primers/probes. Ct values were transformed to copies per ml of wastewater using standard curves for N1 and N2 targets established with synthetic SARS-CoV-2 RNA (Twist Bioscience) as the template. We averaged the viral concentration data on the same day in the northern and southern influents and then multiplied by the daily average flow rate to compute the total viral load in the sewershed.

### 2.2. Clinical data source

The clinical COVID-19 case data for Norfolk, Suffolk, and Middlesex Counties served by the Massachusetts wastewater treatment plant were downloaded from Massachusetts government website (www.mass.gov). We summed the number of clinical cases from each county to represent the total cases in the catchment of the wastewater treatment plant, which is used to compare with the modeling results. Temporal fecal viral shedding data from COVID-19 patients were kindly provided by (Wölfel et al., 2020).

### 2.3. Relationship between wastewater viral concentrations and infectious cases

Assuming we can obtain the fecal viral shedding distribution function over time, we can approximate a constant rate of fecal viral shedding over the duration of infectiousness. In this way, the viral RNA production is proportional to the number of people in the infectious compartment *I* of the SEIR model. That is:

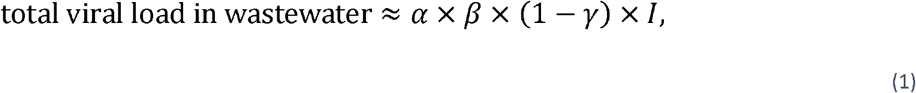

where the proportional constant is defined based on biological parameters similar to (Saththasivam et al. 2021): *α* is the fecal load with unit g/day/person, *β* is the viral shedding rate in stool with unit viral copies/g, and *γ* is the fraction of viral loss in the sewer.

### 2.4. Approximation of fecal viral shedding profile

A key component of this approach is the generation of fecal viral shedding profile. Let *f*(*t*) be the function that describes the temporal fecal viral shedding profile. Upon infection, the shedding of virus in stool should be very small, then reaches a peak before decreasing to 0. Mathematically, this means *f*(0) = 0, lim_*t*→∞_ *f*(*t*) = 0 and *f*(*t*) has a unique maximum for some *t* > 0. While beta and gamma functions are often used to represent *f*(*t*) (Wu et al., 2022a; Ferretti et al., 2020; He et al., 2020), we introduce a phenomenological function *f*(*t*) that is more tractable than the standard beta and gamma functions:

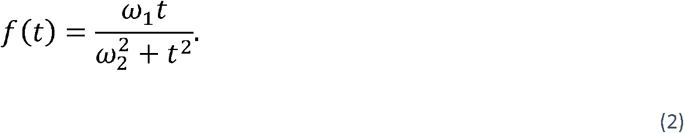

In this form, *ω*_1_ is a magnitude modifier parameter (log_10_ *viral RNA copy per g per day*) and *ω*_2_ (*day*)represents the timing for peak viral shedding and influences the timing and the magnitude of the peak of the viral shedding profile. Specifically, *f*(*t*) peaks at 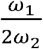 when *t* = *ω*_2_. Thus, if the peak timing and magnitude of the viral shedding profile are known, then can be uniquely defined. It is necessary to mention that *f*(*t*) is the overall viral shedding into the wastewater from infected individuals; however, it mostly means fecal shedding in this work. We did not include the viral shedding from urine or other sources (sputum or saliva) because previous studies showed that no or low level of virus was detected in urine samples of typical patients despite high viral load (Wölfel et al., 2020; Jones et al., 2020), and the total amount of virus in sputum or saliva are likely to be insignificant compared to stool due to the huge difference in volume.

### 2.5. Simple wastewater epidemiological model

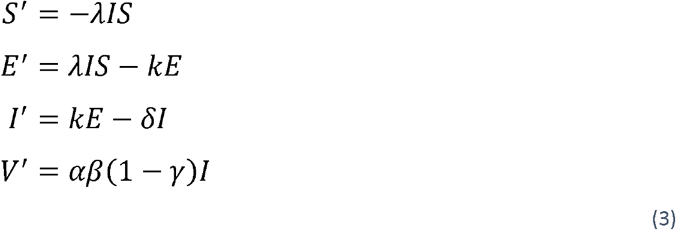

In this model, *S* denotes the susceptible population, *E* is the infected but yet to be infectious population, or the exposed class, *I* is the infectious class, and *V* is the cumulative viral load in wastewater. The *R* compartment (recovered individuals) does not contribute to the transmission dynamics in the SEIR model, hence omitted here. Susceptible people are infected by the infectious class at a rate *λI*. Exposed individuals become infectious at a rate *k*. Infectious individuals recover at a rate *δ* and shed virus at a rate *α* × *δ*, where is the fecal load and *δ* is the average viral shedding rate in Eq (1). is the viral degradation and loss rate in the sewer pipes, so only a fraction (1 − *γ*) of virus is detected in the wastewater sample. The expression for *V* follows directly from Eq (1).

Several studies note that infectious virus is detectable in nose and throat swabs only when the total viral load is above 10^5−6^ copies/mL (Killingley et al., 2022, Ke et al., 2021, Wölfel et al., 2020, Kampen et al., 2021). Since certain level of infectious viruses is required for disease transmission, this implies that the infectious period does not start until the viral load (within host) reaches above 10^5−6^ virus copies/mL. This agrees with previous observation that viral load above 10^6^ copies/mL is associated with a high probability of transmission (Ke et al., 2021). Together, these observations suggest that in this SEIR epidemic model, we can separate the exposed class (*E*) based on the duration before viral load reaches 10^5−6^ copies/mL, and the infectious class (*I*) based on the duration that viral load stays above 10^5−6^ copies/mL. This results in an incubation period of about 3 days and an infectious period of 8 days based on the viral dynamics profile in the SARS-CoV-2 Human Challenge experiment in healthy young adults (Killingley et al., 2022). These estimates are within previous estimated ranges of 2-7 days for incubation periods (Li et al., 2020, Lauer et al., 2020, Guan et al., 2020) and consistent with the updated guideline from CDC where the average infectious duration is about 2 days before and 8 days after symptom onset (CDC, 2022a). Thus, we fix the exposed duration to 3 days, which is equivalent to fixing 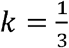 per day (Figure 1A). Similarly, we fix the infectious duration to 8 days, which is equivalent to fixing 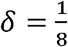 per day. Thus, in our model, parameters *λ, α, β, and γ* need to be estimated.

**Figure 1.**
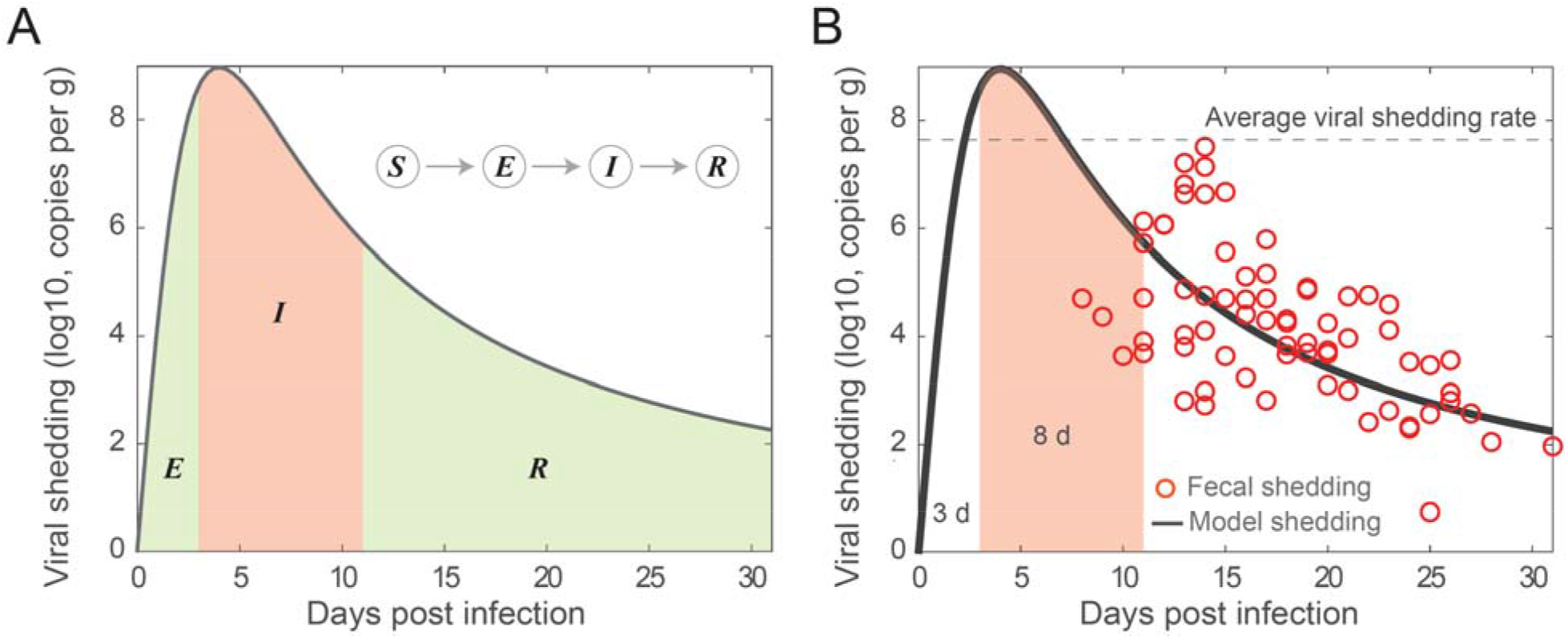
Illustration and fitting fecal viral shedding dynamics. (A) Illustration of the fecal viral shedding dynamics based on the infection progression. The viral shedding profile is divided into three periods shaded: Exposed (*E*), Infectious (*I*), and Recovered (*R*). The red-shaded region is the period of infectiousness *I*, which is corresponding to the compartment *I* in the SEIR model. (B) Fitting of the proposed viral shedding function to viral shedding in hospitalized patients’ infectious period (from day 3 to day 11) is 4.48 × 10^7^ viral RNA per g. The horizontal dash line stool data from (Wolfel et al. 2020). The average viral shedding rate in stool during the is the average fecal viral shedding rate for infectious individuals inferred from the model. The viral shedding peak is at the 4^th^ day post infection.

By fitting the model to wastewater data covering the second wave of the pandemic, specifically, from Oct 2 to Dec 16, 2020, we can approximate the susceptible (to an emerging variant) to be the entire population served by the wastewater treatment plant. For simplification, we assume that there is no infectious individuals initially (*I*(0) = 0), only infected individuals (*E*(0) > 0). The initial value for the virus concentration in wastewater can be taken from the first data point. Thus, *E*(0) is the only unknown initial condition.

The parameters and initial remain to be estimated are: *λ, α, β, γ and E*(0). Since the viral production rate is *αβ*(1 − *γ*), and we only have viral concentration (or total viral load) data, it is impossible to estimate a unique set of values, or specific values, for *α,β, and γ*. For example, the product of *α* = 1, *β* = 2, *γ* = 0.5 is the same as when *α* = 10, *β* = 1, *γ* = 0.9. This reflects the pertinent issue of model identifiability in mathematical models in biology and epidemiology (Tuncer et al., 2022; Eisenberg et al., 2013; Wu et al., 2019; Ciupe and Tuncer, 2022). Thus, an important step in our approach is the direct estimations of *β* and *γ*, which would allow us to identify *α* uniquely. All of the parameters are listed in Table 1.

**Table 1.** Parameters in the model.

|  | Definition | Unit | Value | References |
| --- | --- | --- | --- | --- |
| $S$ | Susceptible population | People | $S(0) = 2.3 \times 10^6$ - fixed | (Wu et al., 2022b) |
| $E$ | Exposed population | People | $E(0)$ – fitting | |
| $I$ | Infectious population | People | $I(0) = 0$ - fixed | |
| $\lambda$ | Transmission rate | Per day per person | fitting | |
| $1/k$ | Exposed duration | Day | 3 days | Wölfel et al., 2020; Killingley et al., 2022; Wu et al., 2022a; Van Kampen et al. 2021; |
| $1/\delta$ | Infectious duration | Day | 8 days | Wölfel et al., 2020, Killingley et al.; 2022, Wu et al., 2022a; Van Kampen et al. 2021 |
| $\alpha$ | Fecal load | Gram | 51-796 g - fitting | Rose et al., 2015 |
| $\beta$ | Viral shedding in stool | Viral RNA copies per gram | fitting | |
| $\gamma$ | Fraction of viral loss in sewer | Per day | Fitting and estimated | |
| $\omega_1$ | Magnitude modifier | $\log_{10}$ viral RNA per g day | fitting | |
| $\omega_2$ | Peak timing for viral shedding | Day | 4 day - fixed | Killingley et al., 2022; Wu et al., 2022a. |

Note that $\beta$ and $\omega_1$ are obtained from fitting to viral shedding data in stool (Wölfel et al., 2020).

### 2.6. Data fitting

Our goal is to fit the SEIR-V model to viral concentration in wastewater data to infer the true number of cases. Then, we compare the predicted number of cases with the daily reported case data. In our model, the variable *V* is the cumulative viral load in wastewater. Thus, the difference of *V* in every 24-hour period reflects the daily measurement data of total virus concentration in wastewater. To reflect this observation, we aim to minimize the sum of square error (*SSE*_*V*_) between these two quantities in our fitting. Hence, our minimization objective is:

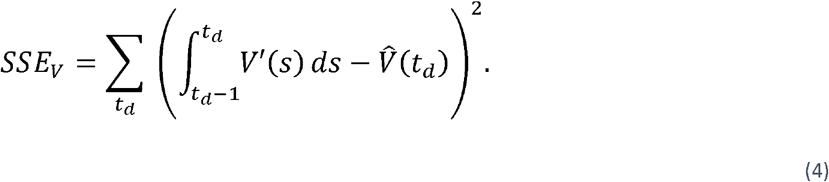

Here, 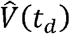 is the total virus concentration experimentally measured on day *t*_*d*_, which equals to viral RNA concentration in wastewater (*C*_*RNA*_)multiplied by the total flow (*F*)data. 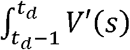 is the corresponding quantity in our model. Once we obtain a reasonable fit to the data, the inferred number of true case is given by:

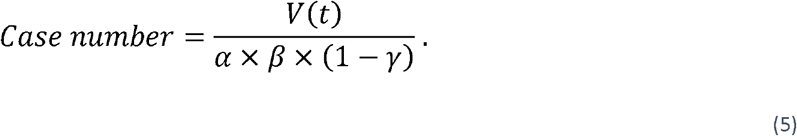

For the minimization algorithm, we use MATLAB function *fmincon* and *multistart*. Similarly, the fecal viral shedding rate function is fitted by minimizing the objective function *SSE*_*f*_:

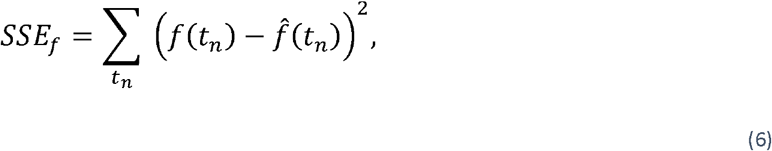

Where 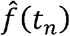 is the fecal shedding data on day.

## 3. Result

### 3.1. Determining the average fecal viral shedding rate in infectious period

We observed that there is a striking similarity in the viral load profiles in nose, throat, and stool for infected individuals from the time of infection to recovery qualitatively (Wölfel et al., 2020, Killingley et al., 2022, Van Kampen et al. 2021). In all three cases, high viral load/shedding is associated with the infectious duration of the infection. This observation suggests that in the classical SEIR epidemic model, we can make the simplifying assumption that the infectious individuals contribute substantially to the viral pools in wastewater. As illustrated in Figure 1A, the viral shedding profile is divided into three periods shaded: Exposed (*E*), Infectious (*I*), and Recovered (*R*). With this framework, we can approximate the viral load in wastewater using the viral shedding from the infectious population. Furthermore, we can estimate the average viral shedding rate based on the viral shedding function *f*(*t*) and the fixed duration of infectiousness (see Materials and Methods).

We fitted the fecal viral shedding function to viral shedding data. Using the best fit parameters, we constructed a fecal viral shedding profile that was used to approximate the fecal viral shedding rate for the infectious individuals. Figure 1B shows the best fit of the model to the fecal viral shedding rate data in Wolfel et al. (Wolfel et al., 2020). Based on the viral dynamics profile in the SARS-CoV-2 Human Challenge experiment in young adults (Killingley et al., 2022), the incubation period (*E*) is about 3 days and the infectious period is about 8 days. We also assumed a five day from infection to symptom onset in the fecal viral shedding data, which is in range of 2-14 days estimated for the general population (CDC, 2022b; Lauer et al., 2020). Furthermore, we fixed the viral peak at day four (*ω*_2_ = 4 *day*). There is no well-established timing of the peak fecal viral shedding rate; however, the peak time for viral load in nose and throat is around 5 days (Killingley et al., 2022) and maybe even earlier in stool (Wu et al., 2022a). The best fit parameter is *ω*_1_ = 71.97 log_10_ *viral RNA copy per g day*. Using the best fit, we estimate the average fecal viral shedding rate for an infectious individual to be:

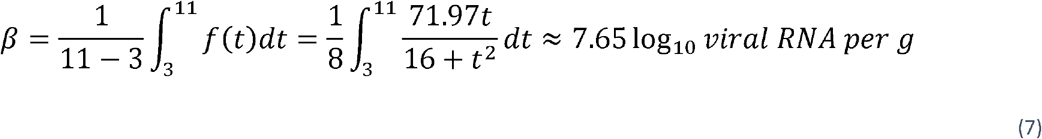

A conversion gives:

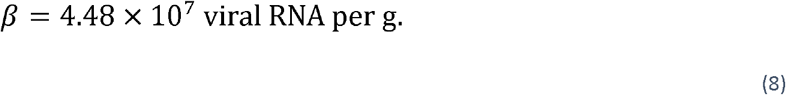

This number is close to the measured median viral RNA load 10^7.68^ (ranging from 10^4.1^–10^10.27^) copies/ml in infected individuals in South Korea (Han et al., 2020), and the extrapolated fecal shedding rate of 10^7.30^ (ranging from 10^5.74^ − 10^8.28^) copies/g of 711 infected individuals in the dormitories at University of Arizona (Schmitz et al., 2021). Thus, we fixed fecal viral production rate in our SEIR-V model to this value.

### 3.2. SEIR-V model captures the temporal dynamics of clinical COVID-19 cases

We developed an SEIR-V model to understand SARS-CoV-2 transmission using WBS data in the second wave of the pandemic and the computed average fecal viral shedding rate during the period of infectiousness. We temporarily ignored the identifiability issue with the conversion equation in an attempt to fit the SEIR-V model to the data. Figure 2 shows the best fit and its inference. We fitted the model to total viral RNA copies in wastewater data up to the grey dashed line (December 16, 2020), then simulated the model out to January 25, 2021, see Figure 2A. The fitting region was chosen before the peak in the viral RNA data, so that we could test the model’s prediction of the peak against the data. Additionally, the fitting region from October 02, 2020 to December 16, 2020 potentially limits the influence from vaccination and the emergence of the alpha variant, which began near the end of 2020.

**Figure 2.**
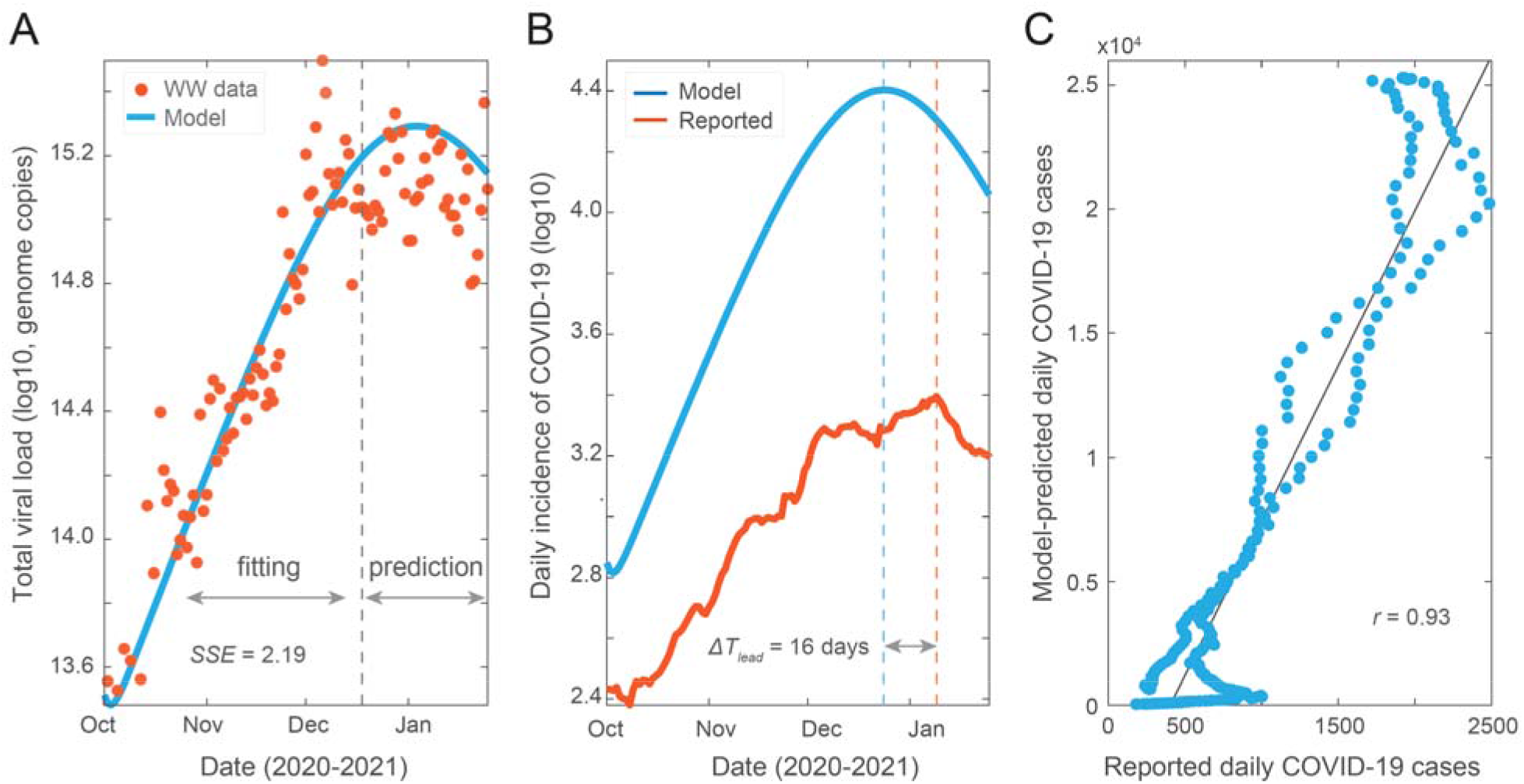
Model fit and prediction to wastewater data covering the second wave of pandemic. (A) Best fit to virus concentration data in wastewater from October 2 to December 16, 2020 (dashed grey line), and model prediction to January 25, 2021. Red dots are the measured viral load in wastewater and blue curve is the modeling result. (B) Model estimation of the true number of COVID-19 cases (blue curve) and clinically reported cases (red curve). The blue and red dashed lines are dates when the two curves peak, and *ΔT*_*lead*_ is the time difference between the two peaks. (C) Correlation between simulation cases and reported cases. Best fit parameters: *λ* =9.66×10^−8^ day^-1^ person^-1^, *α* = 249 *g,γ* = 0.08).

Using the best fit parameters, we computed the number of new cases and compared it to the reported cases. As shown in Figure 2B, the model simulation recapitulates the trend of clinically reported daily new cases and predicts an earlier and higher peak than reported case data by 16 days and 10.2 folds, respectively. We made a correlation plot between the model simulated cases and the reported case data (Figure 2C). The higher predicted number of cases and the high correlation coefficient (*R* = 0.93,*R*^2^ = 0.87) imply that the model accurately captures the trend of the reported case data, while accounting for the underreported rate. This indicates that the method preserves both key properties of WBS data, which is that the trend of viral concentration in wastewater leads the trend of reported cases and can be used to estimate the true prevalence without being impacted by the underreporting rate.

In the next step, we demonstrate how the effect of temperature on viral loss rate can be incorporated in our framework.

### 3.3. Incorporation of wastewater temperature improves model prediction

SARS-CoV-2 RNA in wastewater is subject to degradation which is affected by many factors such as temperature and travel time (Bivins et al., 2020a; McCall et al., 2022). We accounted for these factors to determine an approximate value of *γ*, the fraction of viral decay in the sewershed. The daily viral degradation rate in wastewater is described with the Arrhenius equation:

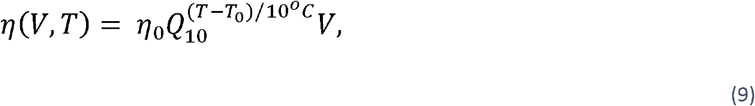

where *η*_0_ is the viral genome degradation rate at ambient temperature *T*_0_ and *Q*_0_ is the temperature dependent rate of change (McMahan et al., 2021; Hart and Halden, 2020). Bivins and colleagues determined that, for wastewater inoculated with high titer at *T*_0_ = 20°C, the mean first-order decay rate of SARS-CoV-2 RNA is *η*_0_ = .0.67 per day (Bivins et al., 2020). Furthermore, *Q*_0_ is typically between 2 and 3 for biological systems, and assumed here to be 2.5 (Běhrádek, 1930; Reyes et al., 2008). Given the relatively constant temperature from October 2 to December 16, 2020 (Figure S1), we used the average temperature of wastewater for the north and south systems for demonstrative purpose, and thus fix *T* = 18°C.

We used the simple exponential decay equation *V′* = −*η*(*V,T*) to estimate *γ*. Let 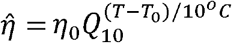, then solving 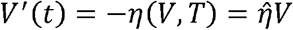 gives:

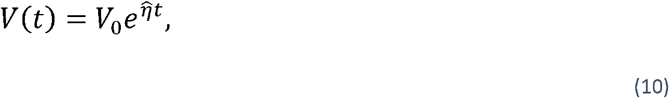

where *V*_0_ is the amount of viral RNA in the sewers at time *t* = 0. Thus, the amount of virus that arrives to the wastewater treatment plant is

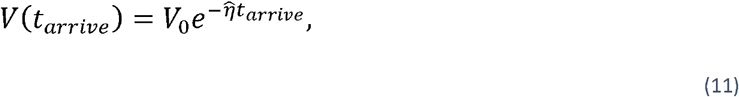

where *t*_*arrive*_ is the time it takes the viral RNA to travel to the wastewater treatment plant after excretion. The time *t*_*arrive*_ includes two parts: the travel time to local sewer pipes and the travel time in the interceptor pipes. Precise estimation of *t*_*arrive*_ is challenging given the varied flow rates and geographical distances to the wastewater treatment plant. Here, we assumed the average travel time is 18 hours. The amount of virus lost is given by *V*_0_ − *V*(*t*_*arrive*_). Thus, the proportion of viral RNA lost in the sewer is given by

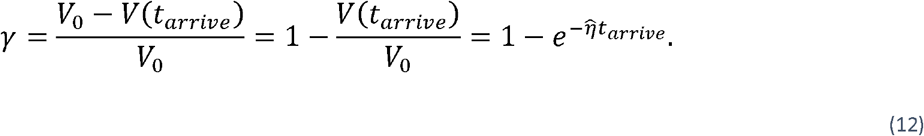

Where the last equality follows from Eq (11). In this case, our calculation yielded *γ* ≈ 0.35, which is in the ranges of previous estimations (Bivins et al., 2020; McCall et al., 2022; Hart and Halden, 2020).

By incorporating temperature effect, the model captures the trend of clinical data with a smaller *SSE*, which is statistically significant based on the corrected Akaike information criterion (Figure 3A, B and S2) (Burnham and Anderson, 2004). We observe that the model simulation predicts an earlier peak than reported case data by 16 days, which is the same as the model without temperature effect (Figure 3B and S2A). Additionally, the model predicts the true number of cases to be about 8.6 times higher than the reported number of cases as compared to a predicted factor of 10.2 without temperature effect (Figure 3B and S2A). The predicted initial exposed population is 2092 people, which is a more reasonable estimate compared to the 11 exposed individuals predicted without temperature (Figure 3B). Those results have shown that incorporating the travel time and temperature reduces the possibility of model unidentifiability and significantly improve the model performance as well as its robustness.

**Figure 3.**
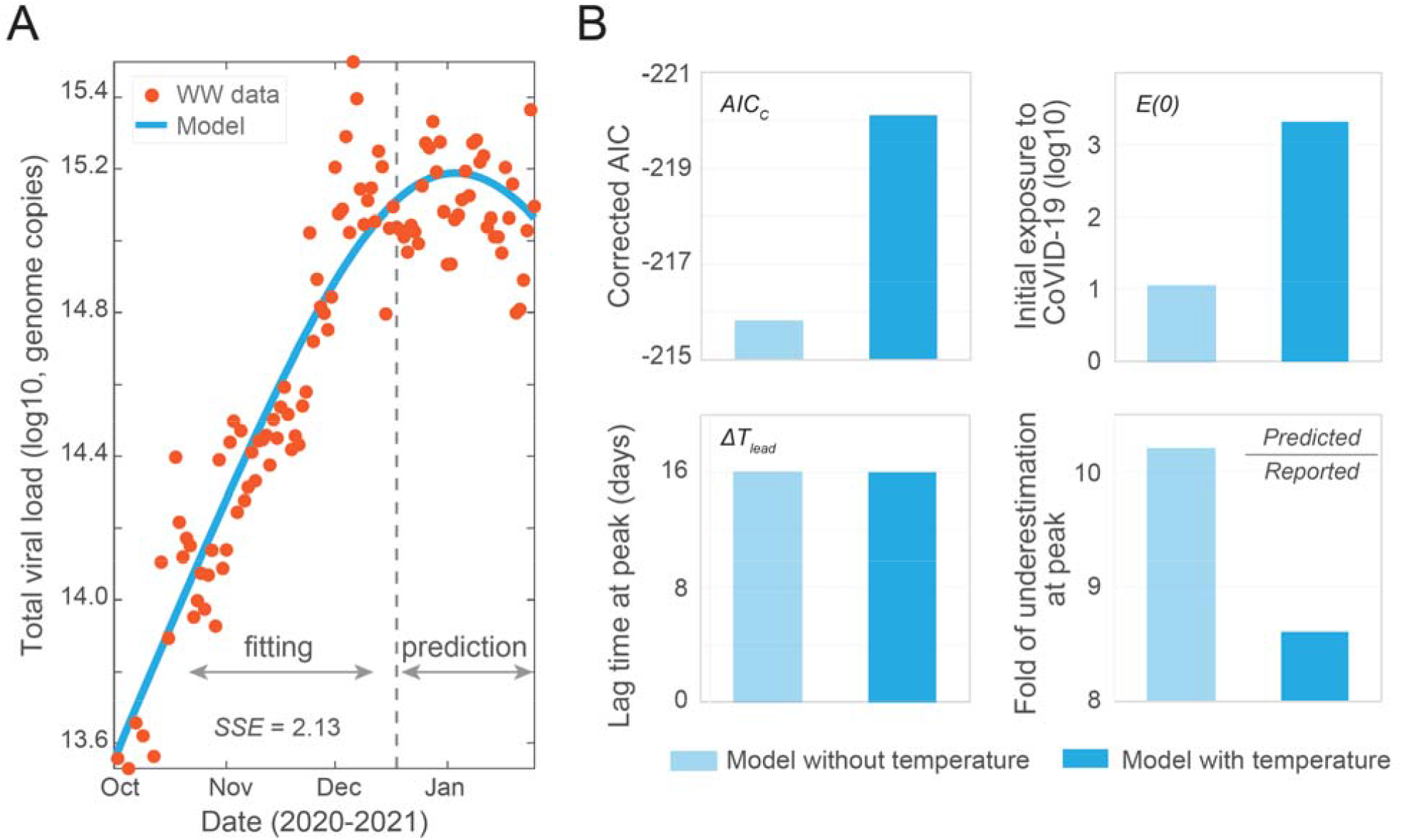
Incorporating temperature effect in the SEIR-V model. (A) Best fit to viral concentration data in wastewater from October 2 to December 16, 2020 (dashed grey line), and model prediction to January 25, 2021. Red dots are the measured viral load in wastewater and blue curve is the modeling result. (B) Comparison of the SEIR-V models with and without incorporating temperature effect. Top left: corrected Akaike information criterion (*AICc*) values, the statistically significant *AICc* difference is 4.3; Top right: initial populations exposed to SARS-CoV-2; Bottom left: wastewater lead time difference at peak, both of the *ΔT*_*lead*_ are 16 days; Bottom right: fold of difference between the number of predicted cases and clinically reported cases. Light blue represents the model without including temperate effect, while blue represents the model with temperature effect. Best fit parameters when incorporating temperature: *λ* = 9.13 ×10^−8^ day^-1^ person^-1^, *α* = 324 *g*, and *E*(0) = 2092 people.

## 4. Discussion

Wastewater collates viral signals excreted by infected individuals across the whole spectrum of disease symptoms from asymptomatic and subclinical-symptomatic to symptomatic (Lee et al., 2020). This inclusiveness of all virus-shedding individuals offers an opportunity to better estimate the magnitude of viral infections in communities (Hart and Halden, 2020; Sanjuán and Domingo-Calap, 2021; Wu et al., 2020). However, it is challenging to convert viral concentrations in wastewater to the number of infected cases. Our group and peers previously reported methods to estimate the infection prevalence by wastewater vial load (McMahan et al., 2021; Nourbakhsh et al., 2021; Wu et al., 2020). These efforts, however, are limited because of inconsideration of dynamic viral shedding rates during the disease course and viral degradation in wastewater.

In this study, we established a quantitative framework to estimate the number of infectious COVID-19 cases and predict SARS-CoV-2 transmission through integrating wastewater surveillance data and development of an SEIR-V model. As an analogy to the four compartments of the SEIR model to simulate infectious disease dynamics at the population level, the individual-level fecal viral shedding course was divided into three periods including exposed (incubation), infectious, and recovery (Figure 1A). The division is based on the observation that the temporal viral profiles in nose, mouth, and stool are strikingly similar qualitatively with high viral load associated with infectiousness (Killingley et al., 2022; Wolfel et al., 2020). In addition, the infectiousness of SARS-CoV-2 is associated with high viral load as reported by multiple studies (Killingley et al., 2022, Ke et al., 2021, Wolfel et al., 2020, Kampen et al., 2021). With this concept, we estimated the population-level average viral shedding rate during the infectious phase using clinical reported SARS-CoV-2 concentrations in hospitalized patients’ stool samples (Figure 1B). This estimated viral shedding rate is an average of infected individuals in the population and does not consider the heterogenous viral shedding dynamics among infected individuals (Wölfel et al., 2020; Killingley et al., 2022; Stanca and Tuncer, 2022). Thus, our model can be improved by feeding viral shedding data during the early phase of the infection and large-scale individual-level shedding dynamics data.

It is noteworthy to mention that the “*I*” in the SEIR model is the “infectious” class, not the “infected” class. Hence, using the viral shedding rate in the infectious period, instead of in the whole shedding period, improves the accuracy of the SEIR model. This contrasts with the conventional approaches that use mean or median viral shedding rate in a group of tested samples regardless of the phase of the infection (Saththasivam et al., 2021; Petala et al., 2022; Schimitz et al., 2021). By focusing on the infectious population, which is also the main contributor of viral shedding in wastewater, we greatly simplify the typical complex structure of the SEIR-type models that implement WBS (Figure S3) and reduces the likelihood of model unidentifiability.

By fitting an SEIR-V model to wastewater data within our framework, we show that the method retains key advantages of using wastewater. Specifically, the inferred case data from the best fit parameters leads the reported case data by 16 days and implies a large ratio (8.6) of true prevalence to clinically reported cases, which are consistent with previous results (Wu et al., 2020; Wu et al., 2022a; Eikenberry et al., 2020; Angulo et al., 2021). We also incorporate the important effect of temperature on the viral degradation rate in a simple manner that is applicable to a larger time scale. We note that extension to incorporate time-dependent variations of the fecal viral shedding rate within this framework is straightforward, but will require careful consideration of the convergence of the numerical method. Together, our work shows the potential and flexibility of the framework to incorporate WBS in epidemic models.

The foundation of our framework is independent of the epidemic model formulation, yet its application depends greatly on the epidemic models for specific situations. For example, if we want to apply the framework to capture a period with significant changes to social behavior, perhaps due to the effect of a social intervention, then an appropriate change to the structure of the SEIR model to reflect these structures is necessary (Johnston and Pell, 2020; Fenichel et al., 2011; Pell et al., 2018). However, if multiple variants are of interest, then the SEIR model itself needs to be extended to a multi-variant version and incorporate known biological properties of different variants (Dyson et al., 2021; Gonzalez-Parra et al., 2021). Similarly, interventions (such as vaccination) and the impact of social gatherings must first be included in the epidemic model prior to its integration within our framework (Saad-Roy et al., 2021; Giordano et al., 2021; Buckner et al., 2021; Makhoul et al., 2020).

Dynamical epidemic models are useful tools to track the pandemic progression and to assess the potential impact of hypothetical situations such as the stay-at-home order or the emergence of a resistant viral strain. However, sparsely reported case data with high uncertainty, due partially to the high underreporting rate, can compromise the ability of epidemic models to provide an accurate forecast of the pandemic and limit their application to retrospective studies. Hence, WBS, which bypasses both the tremendous difficulty in data collection faced by the standard clinical reporting practice and the high underreporting rate, represents a potential solution to address this challenge faced by the modeling community. WBS data also provides a leading indicator of the pandemic progression and is not limited to SARS-CoV-2, thus it can further enhance the prediction and applicability of epidemic models. Together, this aspect of our framework highlights the importance of interdisciplinary collaboration to better address public health concerns

## 5. Conclusions

In this study, we have established a quantitative framework to estimate COVID-19 prevalence and predict SARS-CoV-2 transmission by incorporating WBS data in a simple epidemic SEIR-V model. The main conclusions are:

- We constructed a simple and effective framework to incorporate WBS data to epidemic models. The developed SEIR-V model captures the temporal dynamics of clinical COVID-19 cases and preserves key advantages of WBS data over reported case data.
- We illustrated how the effect of travel time and temperature on viral decay can be incorporated within our framework to improve model performance and robustness, which is an important component to model disease transmission in real world application.
- The modeling framework is a valuable platform to integrate WBS with epidemic models to provide accurate and robust estimates of the pandemic progression and examine the potential impact of interventions to inform public health decision making.

## Data Availability

All data and code produced in the present study are available upon reasonable request to the authors.

## Acknowledgement

This work is supported by Faculty Startup funding from the Center of Infectious Diseases at UTHealth, the UT system Rising STARs award, and the Texas Epidemic Public Health Institute (TEPHI) to F.W. This work was also supported by Director’s postdoctoral fellowship at Los Alamos National Laboratory to T.P.; Y.K. and S.B. are partially supported by the US National Science Foundation Rules of Life program DEB -1930728 and the NIH grant 5R01GM131405-02.

## Supplementary Figures

**Figure S1.**
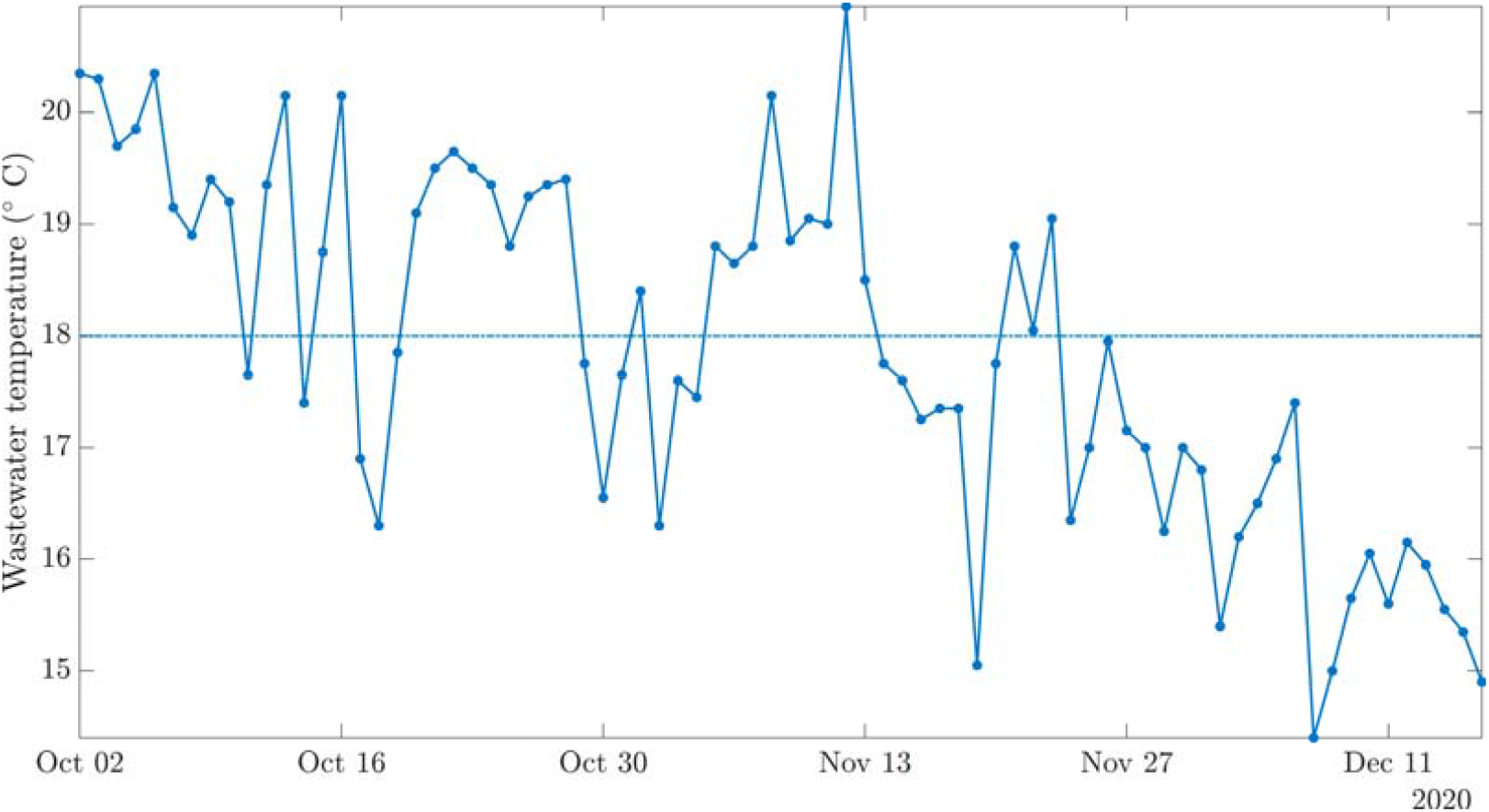
Wastewater temperature data during the model fitting period. Over the fitting duration (from Oct 02 to Dec 16, 2020), the average temperature is relatively constant at around 18 degrees Celsius.

**Figure S2.**
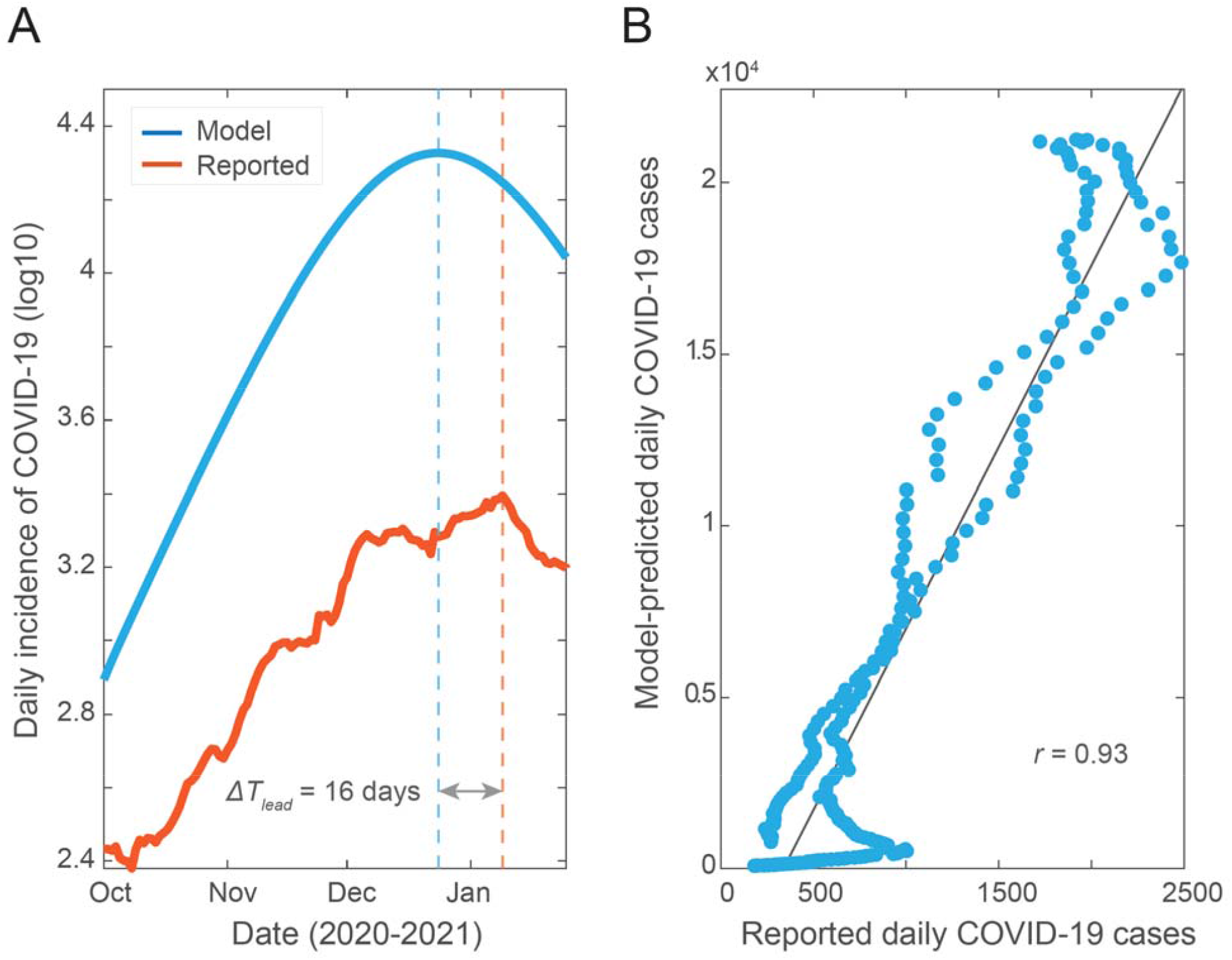
Predicted cases and correlation plot for the SEIR-V model with temperature effect. The wastewater data covers the second wave of pandemic (October 2, 2020 to January 25, 2021). (A) Model simulation of the true number of COVID-19 cases and clinically reported cases. (B) Correlation between the number of model-predicted cases and reported cases (daily).

**Figure S3.**
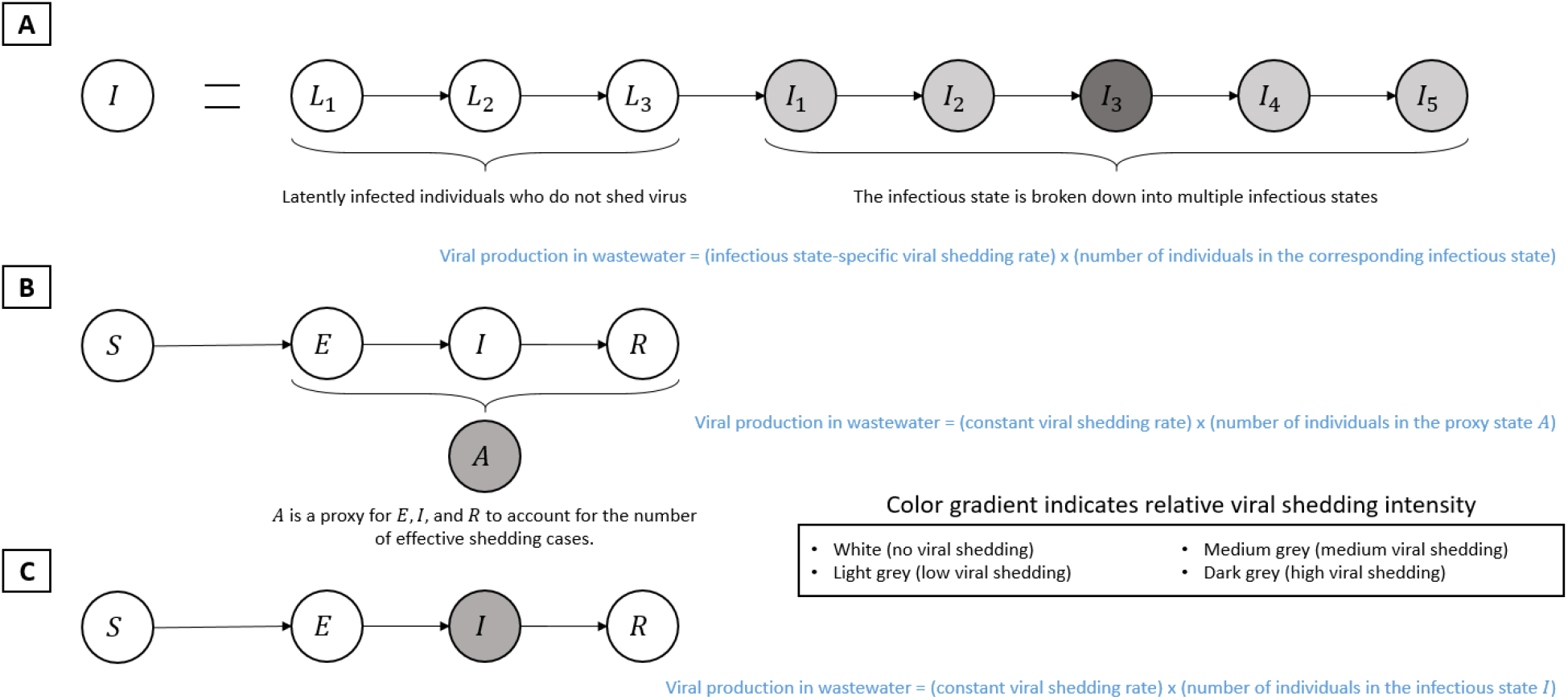
Comparison of underlying frameworks for modeling of wastewater surveillance data. Two general methods have been reported to connect viral concentration in wastewater to epidemic model. (A) is representative of Brouwer et al. and Nourbakhsh et al. (Brouwer et al., 2022; Nourbakhsh et al., 2022), (B) is representative of Proverbio et al. (Proverbio et al., 2022), and (C) is the proposed model framework.

